# Effects of visual arts on persistent pain: A systematic review

**DOI:** 10.1101/2023.05.24.23290506

**Authors:** Wen Hao Matthew Lee, Ali Gholamrezaei, Fereshteh Pourkazemi

## Abstract

**Background:** Persistent pain impacts 30% of people worldwide. Evidence on the effectiveness of visual arts in treating persistent pain seems to be emerging. Thus, the aim of this systematic review was to investigate the impact of visual arts on patients with persistent pain.

**Methods:** Studies were identified by searching seven databases from perception until Jan-2019, then screened by two independent reviewers. Studies were included if they were published controlled trials investigating the impact of visual arts on participants with persistent pain. Studies were excluded if they were abstracts, sampled participants who could not express pain, did not report relevant outcome measures, or did not have a comparator group. The study qualities were assessed by the PEDro scale.

**Results:** After removing duplicates, 2,732 titles and abstracts were screened. Of 125 full-texts, four satisfied the eligibility criteria; all published within the last decade. Three of four controlled studies were randomised-controlled trials. Studies were conducted in inpatient settings (n=2) and outpatient clinics (n=2). Three studies included elderly participants (>60y/o), while one included patients with HIV (>18y/o). Visual arts interventions included painting, drawing, crafting, and others. Two studies utilised visual arts as the sole treatment, while two studies used visual arts as part of a multimodal treatment. Comparators received usual care in two studies, a music intervention in one, and an art-therapy video in another. The common outcome measure in all studies was pain level (0-to-10 scale). All studies also included psychosocial outcome measures. Quality of studies ranged from grade four to eight on PEDro scale; two had “high” quality, and two had “fair” quality. All studies reported statistically significant improvements in pain within intervention groups.

**Conclusions:** Visual arts seem to benefit patients with persistent pain. Further investigation on the clinical significance of these positive findings on pain and other biopsychosocial factors are required.

## Introduction

### The problem of pain

Pain is a universal human experience that can potentially become a debilitating impairment if not addressed adequately. Globally, around 30% of people suffer from persistent pain; the statistics are uncorrelated to countries’ Human Development Index (Elzahaf et al., 2012), implying that this health problem is prevalent in communities regardless of the healthcare system or wealth status of a nation. One in five Australian patients (19.2%) experience persistent pain, with 86.1% of them using medications for pain management (Henderson et al., 2013). Pain commonly impacts patients’ quality of life and poses as a significant economic burden to individuals and the public health system. The economic burden of persistent pain in Australia is estimated to be $34.3 billion per year ($10,847 per persistent pain patient) (Economics, 2007).

### Definition of pain

Pain is defined as “an unpleasant sensory and emotional experience associated with actual or potential tissue damage or described in terms of such damage” (Merskey, 1979). It is a complex, subjective experience that can be influenced by various physiological factors (Melzack, 1999). Hence, there are several theories attempting to explain pain. Traditionally, nociception – the nervous system’s response to potentially harmful stimuli – was thought to be the main origin of pain (Woodworth & Sherrington, 1904). Nociceptor activity would influence behaviour and movement to prevent or limit physical injury (Baliki & Apkarian, 2015). Acute pain is usually indicative of tissue damage or potential tissue damage, serving as a protective mechanism to prevent further damage, especially during the early stages of tissue healing. Though acute pain perception could be temporarily modulated by one’s perception of environmental threats and rewards (Baliki & Apkarian, 2015), it should be expected to improve proportionally to the rate of tissue healing, and resolve within three months of injury. Conversely, persistent pain is not indicative of actual tissue damage and hence, lasts beyond three to six months of injury or beyond the tissue healing time (Sluka, 2016). Persistent pain development is associated with changes in the peripheral nervous system (e.g. reorganisation of receptors and sensory neurones) and central nervous system (e.g. neural reorganisation within the spinal cord and/or brain) (Apkarian et al., 2011; Baliki & Apkarian, 2015). Changes in the peripheral nervous system can sensitise these neural structures and lead to a pain experience that is irrespective to nociception and tissue damage. Changes in the sensory and motor cortexes of the brain can often lead to distorted body image and cortical representation, altering the subjective experience of pain and other forms of sensory-motor feedback (Senkowski & Heinz, 2016; Tsay et al., 2015). Furthermore, persistent pain can potentially be influenced by other factors such as damage to neurological tissues, referred pain, and psychological factors such as emotional amplification (Vachon-Presseau et al., 2016), depression, anxiety, and post-traumatic stress disorder. Therefore, failure to address this multifaceted impairment could potentially lead to adverse consequences.

### Common pain management strategies

The choice of most appropriate form of pain management/treatment(s) depends on the stages of healing, chronicity, and other psychosocial factors. Pain management can include passive treatments (e.g. medications, surgeries, manual therapy, physiotherapy electro-physical modalities) and/or active treatments (e.g. exercise, psychotherapy, psychology). The use of passive treatments does not involve much patient participation, possibly leading to unhealthy expectations about pain-relief, overreliance and reduced activity (increased disability) (Haggan, 2018). Furthermore, treatments vary in short-term and long-term pain-relieving effects, and side effects. Medications for pain include non-opioids (e.g. paracetamol, NSAIDs), opioids, and adjuvants (anti-depressant, neuroleptics, anti-convulsants), prescribed with reference to the World Health Organisation guidelines (Stjernsward, 1986). A third of patients with pain depend on opioids, which have a wide range of psychological and physiological side effects (Ricardo Buenaventura et al., 2008), to control chronic non-cancer pain (Furlan et al., 2006). Sometimes long-term opioid use might even be inappropriately prescribed (Holliday et al., 2013), putting the patient at risk of addiction and side effects without the intended pain-relief. The risks associated with passive interventions such as surgery and long-term medication use warrant a search for other non-invasive and active pain-relief options.

### Active treatments for persistent pain

Active treatments require patients to actively participate in their recovery. Exercise-related therapy and physical activity are examples of active treatments for persistent pain. An overview of systematic reviews reported that exercise and physical activity did not worsen persistent pain and led to a statistically and clinically insignificant decrease in persistent pain (Geneen et al., 2017). Furthermore, there was a significant small-to-moderate improvement in physical function and disability, without negative effects on psychological health and quality of life (Geneen et al., 2017); unlike passive treatments that do not improve physical function and/or come with harmful psychological and physiological side effects. At times, active and passive treatments could collectively lead to better pain relief, as seen in knee osteoarthritis pain where such a combination is more effective than either an active or a passive treatment alone (Jansen et al., 2011).

Recently, new active approaches of pain management such as graded motor imagery (GMI) have been developed, targeting the central changes associated with persistent pain. GMI generally consists of laterality training (distinguishing between left and right limbs), imagined motor movements (visualising pain-free movements of the painful body part), and mirror therapy (moving the unaffected side in the mirror to create the visual illusion that the painful area is moving without pain) (Priganc & Stralka, 2011). Full GMI programmes have shown to have a large pain-relief effect size compared to usual physiotherapy (Bowering et al., 2013), and even lead to decreased level of pain-associated disability (Moseley, 2004, 2006). The effectiveness of GMI could be linked to the reorganisation of the somatosensory cortex as seen in fMRI changes (Walz et al., 2013).

### Psychosocial impacts of persistent pain

The multifaceted complexities of pain are not limited to physical changes. The emotional affective and cognitive behavioural aspects significantly impact the experience of pain. Patients with persistent pain are often affected by the feelings of depression (Ong & Keng, 2003) and anxiety sensitivity (Ocanez et al., 2010), which might further accentuate their pain experience. It is evident that persistent pain is closely co-related to poorer psychological functioning and heightened psychological distress, possibly increasing the patient’s physical and emotional sensitivity to pain (Burke et al., 2015). This can develop into a vicious cycle of persistent pain, heightened sensitivity to pain, worse experience of persistent pain, and even higher sensitivity to pain (with decreased psychological function). These factors could ultimately lead to undesirable consequences to the individual and society. For example, suicidal behaviour is two to three times more prevalent in persistent pain patients than the worldwide average (Campbell et al., 2015). The common co-occurrence of persistent pain and psychological problems make it difficult to treat (Velly & Mohit, 2018), especially if rehabilitation is limited to physical treatments. Therefore, holistic treatments for persistent pain should not only focus on the physical aspects of pain, but also the psychosocial impacts of pain.

### Psychological interventions for persistent pain

Psychological interventions such as cognitive-behavioural therapy (CBT), acceptance and commitment therapy (ACT), and mindfulness, can help to deal with persistent pain (Hughes et al., 2017; Knoerl et al., 2016; Veehof et al., 2016). These interventions involve the patient actively managing their beliefs and responses to pain and injury. Research suggests that CBT can influence intrinsic brain connectivity in patients experiencing persistent pain, leading to a significant decrease in pain and increase in self-efficacy (Shpaner et al., 2014). ACT has been shown to be significantly more effective than controls in improving pain-related mental health and function, without significantly impacting pain intensity and quality of life (Hughes et al., 2017).

### Creative arts and health

Engaging in creative activities (producing or appreciating art) has been shown to drive neuroplasticity through cortical reorganisation and increased blood flow to associated parts of the brain (Demarin et al., 2016), possibly leading to brain changes similar to that of GMI or other psychological interventions mentioned above. Hence, the arts could improve a person’s perception of pain, overall mental health, and function. Creativity and the arts (including various forms of visual art, music, and dance) has been shown to have potential effects on brain activity, emotional state, physiological markers (such as epinephrine and endorphins) and hence, influence health outcomes (Demarin et al., 2016). Furthermore, art forms that include movement (e.g. dance, drama, painting, sculpting) could lead to improvements in physical function similar to that of exercise and physical activity (as mentioned above).

Several literature reviews have explored the impact of music (Bradt et al., 2016; O’Callaghan, 1996), visual arts (Deshmukh et al., 2018; Ruddy & Milnes, 2005), dance (Bradt et al., 2015; Karkou & Meekums, 2017), and drama (Ruddy & Dent-Brown, 2007) on different physical and psychological health outcomes. Of these different art forms, only music has been shown to have a large positive effect on pain (Bradt et al., 2016; Garza-Villarreal et al., 2017; Lee, 2016). Existing reviews on the impacts of visual arts on health are mainly limited to mental health illnesses such as schizophrenia and dementia (Deshmukh et al., 2018; Ruddy & Milnes, 2005). In a randomised controlled trial, painting has been shown to significantly reduce pain in patients with Alzheimer’s Disease (Pongan et al., 2017). However, no systematic review has explored the impact of visual arts on persistent pain. Ongoing reviews (as registered on PROSPERO) are exploring how visual arts could impact mental health and quality of life; none of them are evaluating the effectiveness of visual arts in improving outcomes for patients with persistent pain.

Therefore, the following systematic review aims to consolidate the available literature to determine the effects of visual arts on persistent pain, filling the gap in the existing research. This could possibly help to guide future clinical practice and pave the way for further research in this area (Oxman et al., 1994). The research question is: How does creation or observation of visual arts impact perception of pain in patients with different types of persistent pain, compared to other pain management interventions?

## Methods

### Study design

In order to address the study’s aim and answer the research question, the systematic review study design was chosen. The goal of a systematic review is to produce consolidated, high quality evidence to answer a particular research question. Systematic reviews allow clinicians, researchers, and policy makers to have access to a high-quality overview of the existing evidence regarding a topic (Ferreira Gonzalez et al., 2011; Mulrow, 1994). This is done by identifying all relevant literature containing empirical evidence that meets the pre-set inclusion and exclusion criteria, thoroughly synthesising the available data, to ultimately fulfil the aim of the study and conclusively answer the research question (Lefebvre et al., 2009).

Although a systematic review is typically difficult to conduct in an emerging field of research such as Art and Health, this research method was chosen over other review methods due to the large volume of literature (of varying quality) found during the preliminary search conducted in MEDLINE, Embase, and PsychINFO (refer to *Appendix 1* for the search strategy used). As such, it would be important to limit the search by only including high-quality controlled trials, whilst excluding low-quality uncontrolled trials or case studies.

The methodology of this systematic review was based on the steps listed in Peat (2001):

The strength of conducting a systematic review is that it seeks to extract all available information regarding an area of research (Grant & Booth, 2009), giving us a good overview of the effectiveness of a treatment (i.e. visual arts) in dealing with a health problem (i.e. persistent pain). Furthermore, running a systematic review is an efficient way to obtain high-quality information (Mulrow, 1994). Other forms of research (e.g. controlled trials, clinical trials) usually involve more funding, putting human test subjects at some level of risk, and time spent on recruitment. Systematic reviews do not involve such risks.

However, compared to other scientific review designs (e.g. scoping review), the systematic review has a relatively restrictive nature (e.g. limiting the search to controlled trials). Hence, it might not give us adequate information about why the intervention is or is not effective (Grant & Booth, 2009). The generalised results of the study might not be applicable to specific patients.

Despite its limitations, given the vast number of articles found during the trial search (over 128,000 articles identified from Embase alone), the search findings had to be restricted to randomised controlled trials (over 15,000 randomised controlled trials identified from Embase alone). Hence, the systematic review was the most appropriate study design for investigating the effects of visual arts on pain.

### Eligibility criteria for the studies

Studies were included if they were published full papers that examined the impact of visual arts on participants (of any age) who had persistent pain. Studies were excluded if they (1) were not published full papers (abstracts or conference proceedings); (2) did not sample participants who could express pain; (3) did not report at least one outcome measure demonstrating the impacts of visual arts; (4) did not include objective measures of pain; or (5) did not have a comparator or control group. The details of inclusion and exclusion criteria can be found in *Appendix 2*.

### Data collection

The data collection for the systematic review was conducted as per Preferred Reporting Items for Systematic Reviews and Meta-Analyses (PRISMA) 2009 guidelines (Moher et al., 2009). Data collection began with finding articles within the scope of study; which included selecting the appropriate databases, developing a search strategy, and systematically screening the search results (Gough et al., 2017). The first author decided on the most appropriate databases and search terms with a University librarian.

### Information Sources

Studies were identified by searching MEDLINE (via OVID), Embase (via OVID), PsycINFO (via OVID), Cochrane, CINAHL, PEDro, and Scopus from perception until present. All papers were thoroughly verified, and papers of all languages corresponding to the selection criteria listed above will be included in the analyses.

### Search

The search strategy was defined according to the main concepts of the review (“visual arts” and “persistent pain”). We used the search terms ‘visual arts’ (and all other subsets of visual arts) combined with pain outcomes. The search strategy was run (as shown in *Appendix 1*) on Ovid MEDLINE. The same search strategy was then adapted and run on Embase, PsychINFO, PEDro, CINAHL, Scopus, and Cochrane from perception until 23 January 2019. Articles from each database was exported to an Endnote library and organised into groups. There were 3,924 records found in total.

### Study Selection

After identifying relevant literature by applying our search strategy in the appropriate databases, studies were imported to COVIDENCE – an online tool developed in 2013 to improve the management of systematic reviews (Babineau, 2014) – which screened for duplicates and recommended the removal of duplicates appropriately.

After the removal of duplicates, the titles and abstracts were screened against the set eligibility criteria and included or excluded accordingly. The full-texts of the remaining articles were screened against the eligibility criteria and the reasons for exclusion were recorded. All the screening and eligibility assessment was performed by two reviewers, independently. Disagreements between reviewers were resolved through face-to-face discussions. Authors of the original studies were contacted for clarifications or any additional data. The number of papers found at each stage of the study selection process (identification of papers, title and abstract screening, full-text eligibility screening, inclusion/exclusion) was recorded on a PRISMA 2009 diagram (as seen in Figure 2).

**Figure 1.**
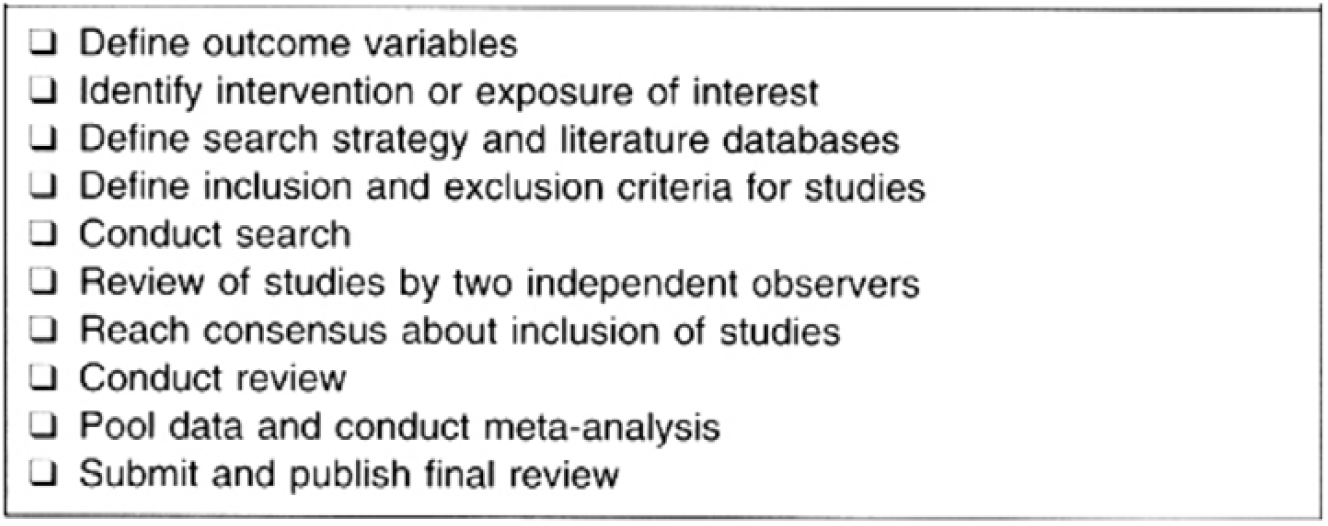
Systematic Review Methodology (Peat, 2001)

**Figure 2.**
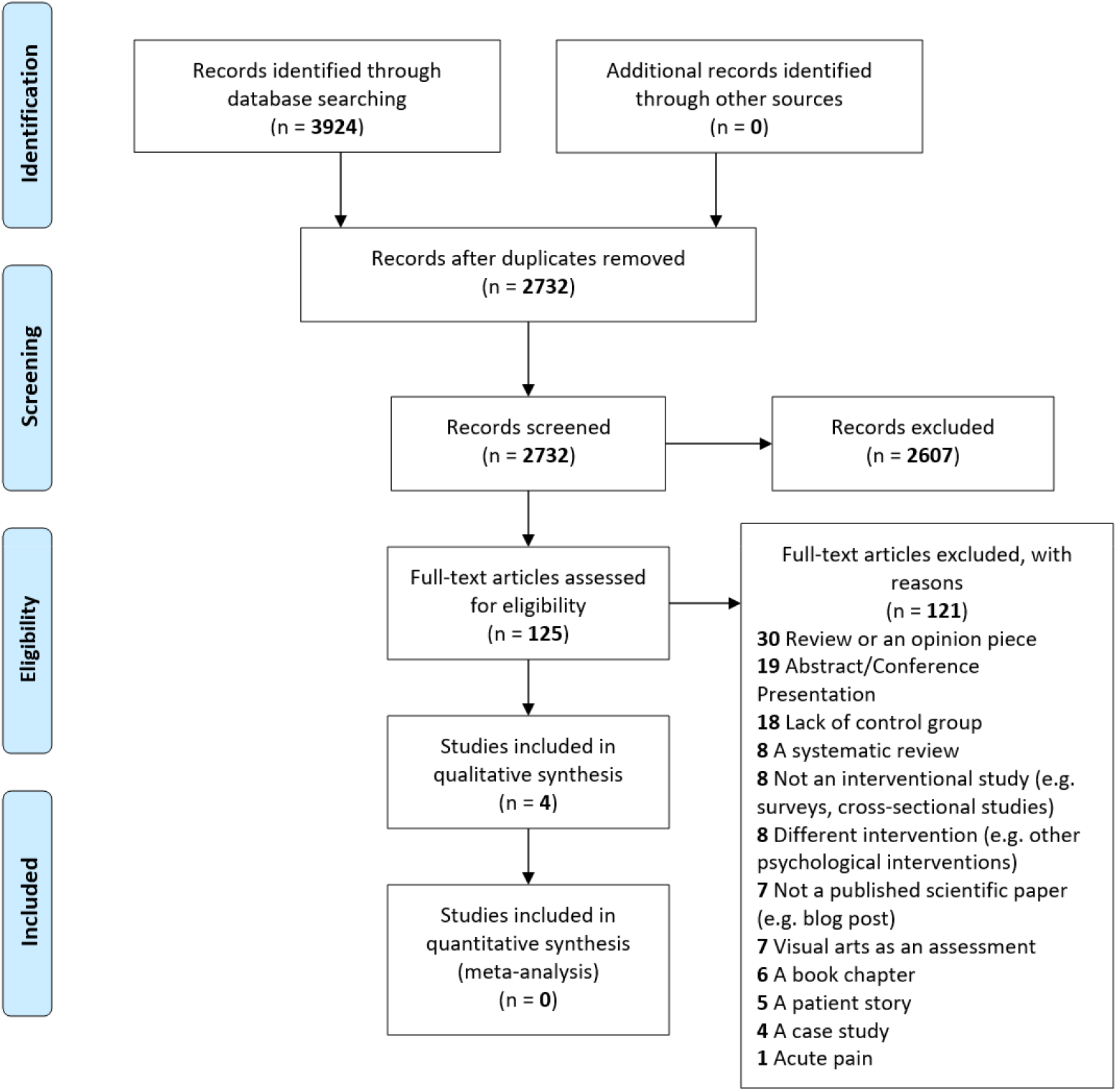
PRISMA 2009 Flow Diagram of Results

### Data collection processes

Data was extracted from all included full-texts. The data included the author’s names, year of publication, country, number of participants, participant demographics, types of persistent pain, types of intervention provided, control, outcome measures, timing of measurements, dropout rates, follow-up details, results, and study quality. Study quality was assessed with the PEDro scale – a 10-point scale used to grade the quality of randomised-controlled trials and their risk of bias (PEDro, 1999; Sherrington et al., 2000). This assessment tool was chosen because it was designed for rating randomised-controlled trials, did not require high levels of expertise, and provided an objective measure of quality (score/10). It was used by the two independent reviewers to grade each study, and disagreements were resolved in face-to-face discussions. All data was extracted, organised, and presented in an electronic data sheet.

### Data analysis

Since the included papers did not have homogenous characteristics (population, intervention, control/comparator, outcome measures), the data in the included studies could not be pooled into a meta-analysis. Therefore, the data from all included studies was synthesised and presented narratively – where important features of each study were explored. Similarities and differences between studies were thoroughly evaluated and reported (Ryan, 2013). For studies reporting significant differences in their primary outcome measures, we calculated the 95% confidence interval (CI) of the effect size using the formula: *95 percent CI = mean difference ± 3 x SD /* √*n*; where difference = difference between the means of both groups, SD = the average standard deviation of both groups, and n = the average number of participants in both groups (Herbert, 2000).

The 95% CIs of statistically significant improvements (with p<0.05) were analysed to determine the clinical significance of the results. A statistically significant difference (p<0.05) was considered to be clinically significant if the 95% CI did not include zero.

## Results

### Study selection

The search strategy identified 3,924 studies. After removing duplicates, the remaining 2,732 titles and abstracts were screened against the eligibility criteria by two independent reviewers. Of 125 full-text studies, four satisfied the eligibility criteria, and were included in this review (Pongan et al., 2017; Rao et al., 2009; Tse & Ho, 2010; Tse et al., 2012). The PRISMA flow diagram for the screening process can be seen in Figure 2 and the list of included studies can be found in Table 1.

**Table 1.** Data Extraction Table

| **Author, Year** | **Participants** | **Setting** | **Intervention** | **Control** | **Dosage** | **Assessment time points** | **Key outcome measures** | **Results** |
| --- | --- | --- | --- | --- | --- | --- | --- | --- |
| Pongan et al. (2017) | N= 59<br>(F:M/39:20)<br>20M, 39F)<br>Age ≥60,<br>Patients with<br>Alzheimer's<br>disease | Memory<br>clinics | N= 28<br>Painting<br>Intervention<br>(PI) | N=31,<br>Singing<br>Intervention<br>(SI) | 1x/week,<br>12<br>weeks | Pre-test,<br>post-test<br>(Week 12),<br>follow-up<br>(Week 16) | Pain: NRS-U,<br>NRS-I,<br>SVS-U,<br>SVS-I, BPI<br>Psychological:<br>STAI<br>GDS | Non-significant decrease in pain<br>in both groups as measured by<br>NRS-I (p= 0.057).<br>No difference between groups<br>for NRS-U and SVS-I.<br>Statistically significant decrease<br>in pain in both groups as<br>measured by SVS-U (p= 0.01)<br>and BPI (p= 0.009).<br>STAI: Significant decrease in<br>anxiety over time in both groups,<br>stronger effect of PI.<br>GDS: Decrease in depression in<br>PI group over time. |
| Rao et al. (2009) | N= 79<br>(F:M/20:59)<br>Age ≥18,<br>Patients with<br>HIV | HIV<br>clinic | N= *not*<br>*reported*<br>Art Therapy | N= *not*<br>*reported*<br>Video about<br>Art<br>Therapy | 1 hour | Pre-test and<br>post-test<br>(one hour<br>later) | Pain: ESAS<br>Psychological:<br>STAI (state<br>portion) | Significant improvement in<br>ESAS score in experimental<br>group compared to control group<br>(b=-5.07, p<0.05).<br>Non-significant improved in<br>STAI score in experimental<br>group compared to control<br>group. |
| Tse & Ho (2010) | N= 141<br>(F:M/109:32)<br>Age ≥60,<br>patients who | Inpatient<br>nursing<br>home | N= 59<br>MSSAC | N= 82<br>Control<br>(regular<br>care) | 1x/week,<br>8 weeks | Pre-test and<br>post-test<br>(Week 8) | Pain: NRS<br>Psychosocial:<br>Chinese<br>psychosocial | Significant decrease in NRS for<br>experimental group (p<0.05).<br>Non-significant improvement in<br>pain in intervention group |
|  | were oriented |  |  |  |  |  | tests | compared to control group (p=0.15).<br>Psychosocial: Significant improvement in SHS, R-UCLA, LSIA, GDS scores compared to control group. |
| Tse et al. (2012) | N= 535<br>(F:M/388:147)<br>Age ≥60,<br>patients who<br>were<br>cognitively<br>intact | Inpatient<br>nursing<br>home | N= 296<br>IPMP | N= 239<br>Control<br>(regular<br>care) | 1x/week,<br>8 weeks | Pre-test and<br>post-test<br>(Week 8) | Pain: VRS<br>Psychosocial<br>wellbeing:<br>Chinese<br>psychosocial<br>tests | Significant decrease in VRS in both groups; intervention group had significantly higher improvement than control group (p < 0.001).<br>Psychosocial: Significant improvement in SHS, R-UCLA, LSIA, and GDS scores in experimental group compared to baseline, and compared to control group. |

BPI: Brief Pain Inventory; Chinese psychosocial tests: The Chinese version of the Subjective Happiness Scale, A Chinese version of the Revised UCLA Loneliness Scale, Life Satisfaction Index-A, Geriatric Depression Scale; ESAS: Edmonton Symptom Assessment Scale; GDS: Geriatric Depression Scale; IPMP: integrated pain management program; LSIA: Life Satisfaction Index-A; MSSAC: multisensory stimulation art and craft appreciation program; NRS: Numerical rating scale (NRS-U: usual pain; NRS-I: worst pain); R-UCLA: Revised UCLA Loneliness Scale; STAI: State Trait Anxiety Inventory; SHS: Subjective Happiness Scale; SVS: Simple Visual Scale (SVS-U: usual pain; SVS-I: worst pain); VRS: Verbal Rating Scales

### Study characteristics

#### Participants and settings

All four papers included adult participants (over the age of 18) who could communicate about their level of pain (Pongan et al., 2017; Rao et al., 2009; Tse & Ho, 2010; Tse et al., 2012), three of which included elderly participants (over the age of 60) (Pongan et al., 2017; Tse & Ho, 2010; Tse et al., 2012). Two studies recruited participants from an outpatient clinical setting (HIV clinic and memory clinic) (Pongan et al., 2017; Rao et al., 2009), while the other two studies recruited participants from an inpatient setting (nursing homes) (Tse & Ho, 2010; Tse et al., 2012). Of the four studies, one was conducted in France (Pongan et al., 2017), one in the USA (Rao et al., 2009), and two in Hong Kong (Tse & Ho, 2010; Tse et al., 2012).

#### Intervention

All included studies had visual arts as part of the intervention (Pongan et al., 2017; Rao et al., 2009; Tse & Ho, 2010; Tse et al., 2012); none of them had visual arts as the sole treatment. Visual arts interventions included painting, colouring, sketching, drawing, crafting (with beads, origami paper, glitter, cups), and making photo albums. Two studies had a meaning-making component to the intervention, wherein the participants would take time to explore the meaning of their artwork (Pongan et al., 2017; Rao et al., 2009). The other two studies had arts and crafts as part of a larger intervention including non-artistic interventions (e.g. relaxation techniques, massage, exercise) (Tse & Ho, 2010; Tse et al., 2012). The interventions were delivered by a range of different professionals. One study had the intervention delivered by a painting teacher and a psychologist (Pongan et al., 2017), one study had the intervention delivered by a certified art therapist (Rao et al., 2009), while the other two had the multi-modal interventions delivered by the researcher and a physiotherapist separately (Tse & Ho, 2010; Tse et al., 2012). Three of the studies had weekly interventions that stretched across an eight to 12-week time period (Pongan et al., 2017; Tse & Ho, 2010; Tse et al., 2012), while one study had only one one-hour intervention session (Rao et al., 2009).

#### Control/Comparison

Due to the inclusion criteria of this review, all included studies had a control or comparison group (Pongan et al., 2017; Rao et al., 2009; Tse & Ho, 2010; Tse et al., 2012). All studies had two groups; one intervention group and one control group. The two studies conducted in nursing homes had the control group receiving regular care only (Tse & Ho, 2010; Tse et al., 2012). One of the studies had a control group watching a one-hour video about the art therapy intervention (Rao et al., 2009), while one of the studies had a music intervention group as its comparator (singing) (Pongan et al., 2017).

#### Outcome measures

All four studies included pain-related outcome measures (Pongan et al., 2017; Rao et al., 2009; Tse & Ho, 2010; Tse et al., 2012); each participant rated their level of pain on a numerical scale of 0 (no pain) to 10 (worst possible pain). This information about pain was either collected as a standalone outcome measure (e.g. Numerical Rating Scale (NRS), Verbal Rating Scale (VRS)) or as part of a larger assessment (e.g. the Edmonton Symptom Assessment System (ESAS)). All studies also included other outcome measures that assessed the psychosocial wellbeing of the participants (e.g. State-Trait Anxiety Inventory (STAI), Subjective Happiness Scale (SHS), Revised UCLA Loneliness Scale (R-UCLA), Life Satisfaction Index-A (LSIA) and Geriatric Depression Scale (GDS)). In all four studies, assessors took pre-test and post-test outcome measures (Pongan et al., 2017; Rao et al., 2009; Tse & Ho, 2010; Tse et al., 2012). However, one study did not report the pre-test and post-test values (Rao et al., 2009). Only one study obtained follow-up outcome measures after four weeks beyond treatment time, presenting slightly longer-term effects of the intervention (Pongan et al., 2017). None of the studies reported any harmful effects from the visual arts interventions.

### Results of individual studies

### Risk of bias

The quality of the included studies ranged from a score of 4-8 on the PEDro scale; two were of high quality (6-10/10) (Pongan et al., 2017; Tse et al., 2012), and two were of fair quality (4-5/10) (Rao et al., 2009; Tse & Ho, 2010). All four studies had specified eligibility criteria, collection of key outcome measure(s), and adequate data analysis of these outcome measures; reflecting low risk of bias in these criteria (Pongan et al., 2017; Rao et al., 2009; Tse & Ho, 2010; Tse et al., 2012). None of the studies had blinding of the therapists or participants. The results and details of the quality assessment can be found in Table 2.

**Table 2.** Risk of bias results

| Author/Year | Pedro Score (/10) | PEDro Criteria |  |  |  |  |  |  |  |  |  |  |
| --- | --- | --- | --- | --- | --- | --- | --- | --- | --- | --- | --- | --- |
|  |  | 1 | 2 | 3 | 4 | 5 | 6 | 7 | 8 | 9 | 10 | 11 |
| Pongan et al. (2017) | 8 | Yes | Yes | Yes | Yes | No | No | Yes | Yes | Yes | Yes | Yes |
| Rao et al. (2009) | 4 | Yes | Yes | No | No | No | No | No | Yes | Yes | Yes | No |
| Tse & Ho (2010) | 5 | Yes | No | No | Yes | No | No | No | Yes | Yes | Yes | Yes |
| Tse et al. (2012) | 6 | Yes | Yes | No | Yes | No | No | No | Yes | Yes | Yes | Yes |

### Quality Assessment Table (PEDro Scale)

1. Eligibility criteria were specified (not counted in final score);
2. Subjects were randomly allocated to groups (in a crossover study, subjects were randomly allocated an order in which treatments were received);
3. Allocation was concealed;
4. The groups were similar at baseline regarding the most important prognostic indicators;
5. There was blinding of all subjects;
6. There was blinding of all therapists who administered the therapy;
7. There was blinding of all assessors who measured at least one key outcome;
8. Measures of at least one key outcome were obtained from more than 85% of the subjects initially allocated to groups;
9. All subjects for whom outcome measures were available received the treatment or control condition as allocated or, where this was not the case, data for at least one key outcome was analysed by “intention to treat”
10. The results of between-group statistical comparisons are reported for at least one key outcome;
11. The study provides both point measures and measures of variability for at least one key outcome.

### Synthesis of results

Using the mean difference, standard deviation and number of participants, the 95% CIs were calculated to investigate the clinical significance of these art intervention on patient outcomes. The 95% CIs for all statistically significant changes (with p<0.05) are reported in Table 3. All included studies reported a statistically significant improvement in at least one key outcome measure of pain (NRS, SVS, BPI, VRS, or ESAS) between pre- and post-test measures (Pongan et al., 2017; Rao et al., 2009; Tse & Ho, 2010; Tse et al., 2012). Of the four studies, two studies had 95% CIs that did not include zero (Tse & Ho, 2010; Tse et al., 2012), one study had 95% CIs that included zero (Pongan et al., 2017), and one study did not provide pre-test and post-test data for the calculation of the 95% CIs (Rao et al., 2009). Two studies reported a statistically significant improvement in pain (as measured by ESAS or VRS) with visual arts interventions compared to control groups (Rao et al., 2009; Tse et al., 2012). Of the two studies, one study had 95% CIs that did not include zero (Tse et al., 2012), while one study did not provide post-test data for the calculation of the 95% CIs (Rao et al., 2009).

**Table 3.** 95% Confidence Intervals for statistically significant findings

| Author, Year | Outcome measure | 95% CI of mean change within Intervention group | 95% CI of mean change within Control group | 95% CI of mean change between groups |
| --- | --- | --- | --- | --- |
| Pongan et al., (2017) | SVS-U (out of 4) | -0.33, 0.67 | -0.04, 0.90 | Missing data |
|  | BPI (out of 10) | -0.40, 1.34 | -0.11, 1.25 |  |
|  | STAI (out of 80) | **1.74, 19.90** | Missing data |  |
|  | GDS (out of 30) | -0.36, 6.03 |  |  |
| Rao et al. (2009) | ESAS (out of 90) | NA | NA | Missing data |
| Tse & Ho (2010) | NRS (out of 10) | **1.15, 2.99** | NA |  |
|  | SHS (out of 28) |  |  | **-4.49, -0.71** |
|  | R-UCLA (out of 80) |  |  | **4.31, 11.17** |
|  | LSIA (out of 18) |  |  | **-2.85, -0.23** |
|  | GDS (out of 15) |  |  | -0.02, 2.46 |
| Tse et al. (2012) | VRS (out of 10) | **1.12, 1.92** | **0.23, 1.15** | **0.19, 1.05** |
|  | SHS (out of 28) | -2.04, -0.16 | NA | -1.96, 0.02 |
|  | R-UCLA (out of 80) | -0.31, 3.29 | NA | -1.04, 2.88 |
|  | LSIA (out of 18) | -1.05, 0.33 | NA | -1.29, 0.13 |
|  | GDS (out of 15) | -0.41, 0.83 | NA | -0.60, 0.74 |

BPI: Brief Pain Inventory; ESAS: Edmonton Symptom Assessment Scale; GDS: Geriatric Depression Scale; LSIA: Life Satisfaction Index-A; NRS: Numerical rating scale; R-UCLA: Revised UCLA Loneliness Scale; STAI: State Trait Anxiety Inventory; SHS: Subjective Happiness Scale; SVS: Simple Visual Scale (SVS-U: usual pain); VRS: Verbal Rating Scales

Three of four studies reported on significant effects of visual arts on psychological symptoms; as measured by STAI scores and other psychological parameters (Happiness, Loneliness, Life Satisfaction, Depression scores) (Pongan et al., 2017; Tse & Ho, 2010; Tse et al., 2012). Two studies reported a statistically significant improvement in at least one psychological measure between pre-test and post-test measures (Pongan et al., 2017; Tse et al., 2012); one study one study had a 95% CI that did not include zero (Pongan et al., 2017), while the other had 95% CIs that included zero (Tse et al., 2012). One study reported a statistically significant improvement in four psychological outcomes with visual arts intervention compared to the control group; three out of four outcomes had 95% CIs that did not include zero.

## Discussion

### Summary of evidence

The aim of this study was to investigate the effects of visual arts of persistent pain. The results of this systematic review indicate that this is a new, emerging area of research with limited controlled trials done to date. All included studies were published within the last decade (Pongan et al., 2017; Rao et al., 2009; Tse & Ho, 2010; Tse et al., 2012). The included studies support the use of visual arts as a safe, non-invasive, and beneficial treatment for the pain and psychological wellbeing of patients with persistent pain. These results are promising, considering the complex, multifaceted nature of persistent pain. All studies reported that visual art interventions led to a statistically significant improvement in pain between pre- and post-test measures (Pongan et al., 2017; Rao et al., 2009; Tse & Ho, 2010; Tse et al., 2012); two studies had clinically significant improvements (with 95% CIs that did not include zero). Of the two studies that reported a statistically significant improvement in pain with visual arts interventions compared to control groups (Rao et al., 2009; Tse et al., 2012), one study had clinically significant improvements (with 95% CIs that did not include zero).

Furthermore, all studies reported on additional effects of visual arts on psychological symptoms; as measured by STAI, SHS, R-UCLA, LSIA, and GDS scores (Pongan et al., 2017; Rao et al., 2009; Tse & Ho, 2010; Tse et al., 2012). Since persistent pain is a complex problem that is often associated with psychological problems and psychiatric co-morbidities (Dominick et al., 2012; McWilliams et al., 2003; Tsang et al., 2008), these findings suggest that apart from reducing pain levels, visual arts can be effective in addressing the psychological factors that are commonly associated with persistent pain.

Existing meta-analyses have explored the effects of musical interventions on patients with persistent pain (Garza-Villarreal et al., 2017; Lee, 2016), with pooled results showing the effectiveness of music as an adjunctive treatment for persistent pain. Although the data in this systematic review could not be pooled in a meta-analysis due to heterogeneity, the included studies support the use of visual arts in treating persistent pain. Current studies that explore the effects of visual arts and persistent pain show positive therapeutic effects that are comparable to that of musical interventions.

In one of the included studies, Pongan et al. (2017) compared the effects of music intervention to painting intervention. Both groups showed statistically significant improvement in pain from baseline. In an existing systematic review, there is low quality evidence for music interventions leading to reduction in pain and anxiety in cancer patients (Bradt et al., 2016). Since the painting intervention had similar effects to the music intervention on persistent pain in this study, it could be inferred that painting was also an effective treatment for persistent pain. Hence, it can serve as a feasible alternative to music therapy, especially in cases where participants have speech and/or hearing impairments.

Two of included studies were based in Hong Kong and had similar methods (two groups, control group receiving usual care, eight-week intervention) and participants who had similar characteristics (nursing home setting, above 60 years old, oriented to time and place) (Tse & Ho, 2010; Tse et al., 2012). However, the interventions differed. Tse et al. (2012) had an additional component of physical training. Since physical training has been shown to reduce pain and depression, and to improve physical function and quality of life (Cooney et al., 2013; Geneen et al., 2017), the results from the two studies could not be pooled to estimate the effect size of the visual arts intervention.

Rao et al. (2009) reported statistically significant improvements in ESAS and STAI in the art therapy group compared to controls, they did not include their pre-test and post-test data values. Therefore, it is difficult to assess the true effect size of the intervention compared to controls. Nevertheless, the findings of the study suggest that participating in one art therapy session leads to larger change in physical symptoms compared to psychological symptoms. However, the longer-term effects of therapy were not assessed in this study. Since the art therapy process usually spans eight to 15 weeks (Regev & Cohen-Yatziv, 2018), the long-term efficacy of art therapy on persistent pain has yet to be explored. If patients are able to make significant improvements after each session, and maintain that change over a period of time, art therapy would be a promising option in the treatment of persistent pain.

The involvement of different professionals in the administering of the art intervention could have affected the efficacy of treatment. For example, a certified art therapist (Rao et al., 2009) would have received a lot more specialised training in administering artistic interventions for health-related outcomes as compared to all the other professions. As mentioned above, the length of intervention could also have affected the efficacy of treatment.

As expected for this type of intervention, none of the studies had blinding of participants and therapists who administered therapy. Although the lack of participant and therapist blinding supposedly indicates a high risk of bias, blinding of visual arts interventions is not possible; the therapists would know the treatments that they administer and the participants would know the treatment that they receive.

The mechanism of action of visual arts on persistent pain remains unclear, however there are several theories or suggestions on how visual arts could reduce persistent pain. A potential mechanism of action for art interventions is that it can help divert the patient’s attention away from the pain (and pain-related symptoms) to the artistic work (Malchiodi, 1998), engaging many parts of the brain (cortex, limbic system, brainstem) in the process (Malchiodi, 2011). Also, it provides patients with an open channel of expression, through which they can communicate their suppressed pain and emotions to others (Aldridge, 1993). Collectively, these can help increase patients’ self-efficacy in pain management, gradually assisting them to overcome their pain in the long term.

Although this study did not aim to report on the mental health outcomes, all included studies reported on the effects of visual arts on psychological parameters. Three studies reported a significant improvement in at least one psychological symptom from baseline (Pongan et al., 2017; Tse & Ho, 2010; Tse et al., 2012), and the other study reported a non-significant improvement in STAI scores compared to the control group (Rao et al., 2009). Specifically, painting was shown to have a strong effect on improving symptoms of depression and anxiety over time (Pongan et al., 2017). Future controlled trials and reviews can also investigate the effects of visual arts on the psychological issues that are associated with persistent pain.

### Limitations

Only one study reported a clinically significantly higher improvement in pain compared to the control group (Tse et al., 2012). Moreover, since the minimal clinically important difference of the NRS is two points or 30% (Farrar et al., 2001), it is still unclear if the treatment effect has true clinical value. More research is required to determine the potential effectiveness of using visual arts interventions to treat persistent pain.

While developing the systematic review inclusion and exclusion criteria, both reviewers had agreed on excluding non-controlled trials in order to maximise the quality of included papers. Consequently, 18 full-texts were excluded from this review due to lack of control group; some of these studies reported the effects of visual arts on persistent pain with pre- and post-test measures. The inclusion of these studies might have allowed us to collect and analyse more relevant data. Hence, the stringent inclusion criteria limited the scope of the review. Future reviews could include non-controlled trials for further analysis of treatment effects.

### Clinical implications

Despite the limitations in current evidence for the efficacy of visual arts in treating persistent pain, the intervention has potential to positively affect clinical practice. It presents patients and clinicians with another active, non-invasive treatment option for persistent pain. Visual arts interventions also provide patients with a new way of reflecting about their pain and communicating their pain, which would be particularly useful for clinician-patient communication and education. Clinicians who would like to administer visual arts interventions might need to attend additional training to efficaciously deliver the interventions.

## Conclusion

The results of this systematic review illustrate the emerging body of evidence for the use of visual arts in the treatment of persistent pain; all included studies were published in the past decade. Based on the results of existing studies, visual arts interventions seem to be a beneficial treatment for patients with persistent pain; all studies reported a statistically significant change in pain levels from pre- to post-test. Furthermore, since none of the studies reported any adverse reactions to visual arts interventions, visual arts present as a safe, non-pharmacological alternative for the treatment of persistent pain in clinical practice. However, there was insufficient evidence to prove the clinical relevance of these positive findings; only one study had a clinically significant improvement in pain compared to control groups, with a 95% confidence interval that did not include zero. The analysis of this systematic review was restricted by the limited quantity and quality of controlled trials conducted in this emerging field. Therefore, further high-quality research into the impacts of visual arts on patients with persistent pain is recommended.

## Data Availability

All data produced in the present study are available upon reasonable request to the authors.

**Appendix 1.** Search Strategy as run on Ovid MEDLINE

| # | Searches | Results |
| --- | --- | --- |
| 1 | ART/ | 7187 |
| 2 | ART THERAPY/ | 1371 |
| 3 | art therap*.mp. | 1797 |
| 4 | visual art*.mp. | 465 |
| 5 | colour therap*.mp. | 4 |
| 6 | color therap*.mp. | 92 |
| 7 | CREATIVITY/ | 6603 |
| 8 | creativ*.mp. | 16894 |
| 9 | medicine in the arts.mp. or Medicine in the Arts/ | 4343 |
| 10 | motion pictures.mp. or Motion Pictures/ | 7701 |
| 11 | Paintings/ | 4780 |
| 12 | pictorial works as topic.mp. or Pictorial Works as Topic/ | 10 |
| 13 | portraits as topic.mp. or Portraits as Topic/ | 1454 |
| 14 | science in the arts.mp. or Science in the Arts/ | 7 |
| 15 | SCULPTURE/ | 1042 |
| 16 | sculpt*.mp. | 4159 |
| 17 | Visual Perception/ | 58846 |
| 18 | visual perception*.mp. | 61086 |
| 19 | Color Perception/ | 15878 |
| 20 | color perception*.mp. | 17598 |
| 21 | drawing*.mp. | 28006 |
| 22 | sketching*.mp. | 222 |
| 23 | colo?ring.mp. | 48683 |
| 24 | mask making.mp. | 5 |
| 25 | Chronic Pain/ | 11033 |
| 26 | chronic pain*.mp. | 32539 |
| 27 | (chronic adj3 pain*).mp. | 50028 |
| 28 | Pain Management/ | 29445 |
| 29 | pain management*.mp. | 40977 |
| 30 | 1 or 2 or 3 or 4 or 5 or 6 or 7 or 8 or 9 or 10 or 11 or 12 or 13 or 14 or 15 or 16 or 17 or 18 or 19 or 20 or 21 or 22 or 23 or 24 | 193798 |
| 31 | 25 or 26 or 27 or 28 or 29 | 83312 |
| 32 | 30 and 31 | 435 |

**Appendix 2.**
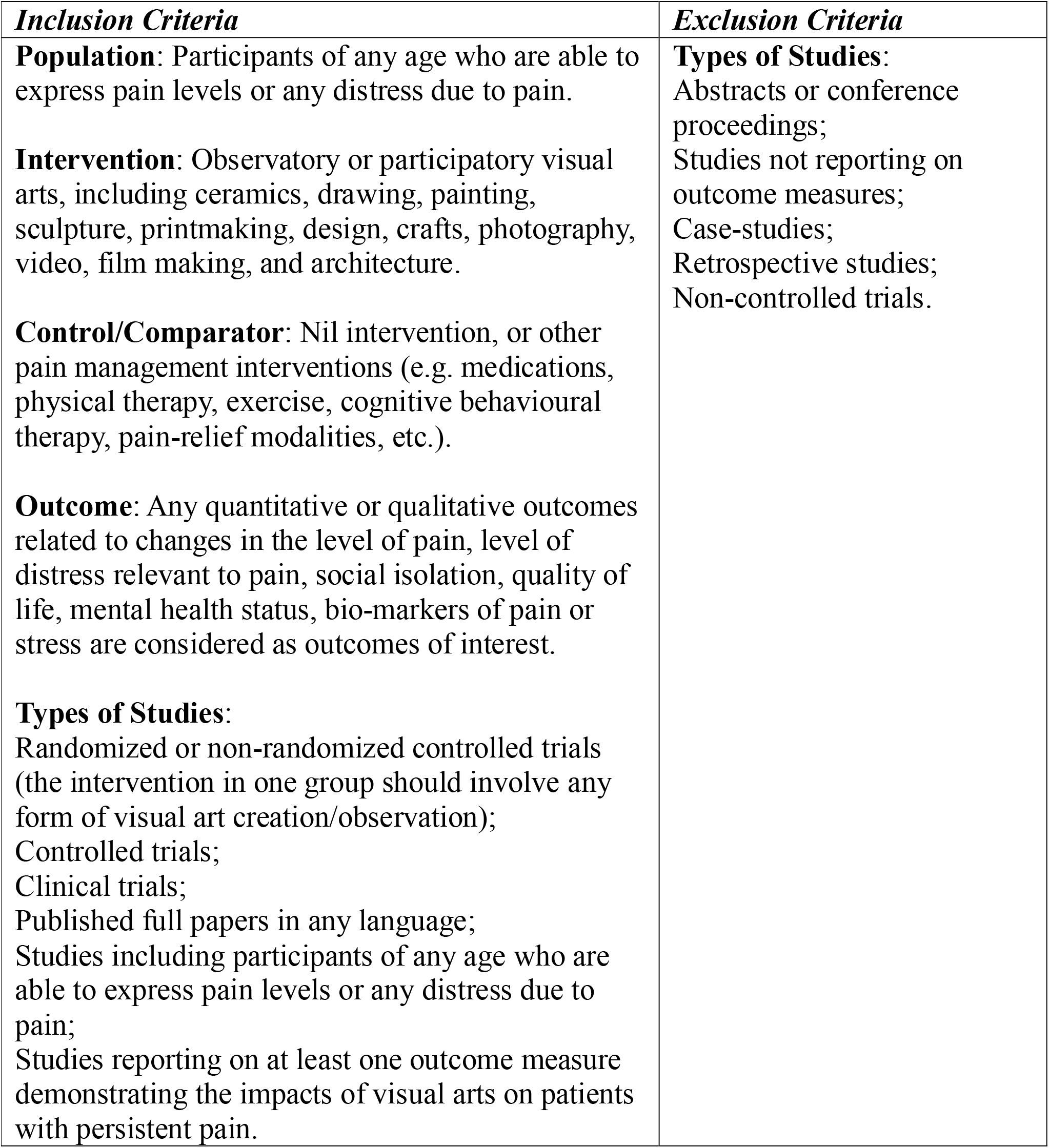
Inclusion and Exclusion Criteria

| ***Inclusion Criteria*** | ***Exclusion Criteria*** |
| --- | --- |
| **Population:** Participants of any age who are able to express pain levels or any distress due to pain.<br>**Intervention:** Observatory or participatory visual arts, including ceramics, drawing, painting, sculpture, printmaking, design, crafts, photography, video, film making, and architecture.<br>**Control/Comparator:** Nil intervention, or other pain management interventions (e.g. medications, physical therapy, exercise, cognitive behavioural therapy, pain-relief modalities, etc.).<br>**Outcome:** Any quantitative or qualitative outcomes related to changes in the level of pain, level of distress relevant to pain, social isolation, quality of life, mental health status, bio-markers of pain or stress are considered as outcomes of interest.<br>**Types of Studies:**<br>Randomized or non-randomized controlled trials (the intervention in one group should involve any form of visual art creation/observation);<br>Controlled trials;<br>Clinical trials;<br>Published full papers in any language;<br>Studies including participants of any age who are able to express pain levels or any distress due to pain;<br>Studies reporting on at least one outcome measure demonstrating the impacts of visual arts on patients with persistent pain. | **Types of Studies:**<br>Abstracts or conference proceedings;<br>Studies not reporting on outcome measures;<br>Case-studies;<br>Retrospective studies;<br>Non-controlled trials. |

